# Fine-Tuning the Llama2 Large Language Model Using Books on the Diagnosis and Treatment of Musculoskeletal System in Physical Therapy

**DOI:** 10.1101/2023.11.23.23298943

**Authors:** Jun-hee Kim

## Abstract

**Backgroud:** Generative language models (GLM) utilize machine learning algorithms to perform various tasks such as text generation, question response, and sentence completion by imitating the language that humans understand and use.

**Purpose:** This study was to fine-tune the Llama2 language model using text data from books on the diagnosis and treatment of musculoskeletal system in physical therapy and compare it to the base model to determine its usability in medical fields.

**Results:** Compared to the base model, the fine-tuned model consistently generated answers specific to the musculoskeletal system diagnosis and treatment, demonstrating improved understanding of the specialized domain.

**Conclusion:** The model fine-tuned for musculoskeletal diagnosis and treatment books provided more detailed information related to musculoskeletal topics, and the use of this fine-tuned model could be helpful in medical education and the acquisition of specialized knowledge.

## INTRODUCTION

In November 2022, OpenAI launched the ChatGPT service based on a generative language model (GLM) called GPT.^1^ This is an important milestone in the development of artificial intelligence (AI) technology, which has brought about major changes in natural language processing and computer science.^1,2^ GLM-based services such as ChatGPT use machine learning algorithms to mimic and generate the language that people understand and use.^1,3^ These GLM-based services can perform a variety of tasks, including text generation, question answering, and sentence completion.^1,3^

GLMs, such as OpenAI’s GPT, Google’s BERT, and Meta’s Llama, are currently being actively developed and have influenced many fields such as economy, industry, and medicine.^4–6^ It is used in information search, customer service, and education, and its impact continues to expand.^7,8^ These GLMs can perform a variety of tasks, including generating text, answering questions, and completing sentences.^9^ The diversity and efficiency of tasks these models can perform depend largely on the amount of information that the models can process and generate based on the data used for training.^10^

However, GLMs are not perfect and still need to be improved. GLMs in service currently cover general knowledge but lack knowledge of specialized domains.^11,12^ This means that the model may have difficulty answering specialized questions accurately.^11,12^ Additionally, these models often fail to provide complete and accurate answers to users’ questions.^13,14^ There is a representative problem called ‘hallucination’, which refers to a phenomenon in which the model generates information that is not in the training data or provides incorrect information.^15,16^ These issues reduce model reliability and can provide incorrect information to users.^15,16^

To solve these problems, there is a need for GLMs tuned to specialized domains. Especially in the medical field, since accurate and reliable information is important, the need for GLMs that include medical information is becoming more and more prominent.^4,12,17,18^ These models should be able to provide medical knowledge to people other than medical experts.^4,19^ These models can also play an important role in efficiently supporting the training of healthcare professionals.^4,20,21^ In particular, it can help students in medical professions understand complex medical terms, basis for medical guidance, and determine appropriate treatment plans for various patient situations.^20–22^

The purpose of this study was to fine-tune the large language model Llama2 with text data from books on the musculoskeletal system in physical therapy and evaluate its usability in specialized areas such as physical therapy, a subcategory of medicine. Therefore, this study compared the answers generated when questions related to the specialized musculoskeletal domain of physical therapy were applied to fine-tuned and untuned models, respectively, to evaluate and confirm the possibility of whether language models fine-tuned to the professional domain can correctly answer professional knowledge.

## METHODS

### Data collection and pre-processing

The process of collecting, processing, and fine-tuning the data to the language model is shown in Figure 1. For this study, books related to the diagnosis and treatment of musculoskeletal disorders in the field of physical therapy were selected. First, the book ‘Functional Anatomy’, which is considered the most basic area in diagnosing or treating musculoskeletal disorders, was selected.^23^ And a book called ‘Kinesiology’, which describes the movement of the musculoskeletal system in the human body, was selected.^24^ In addition, ‘Diagnosis and Treatment of Movement System Impairment Syndromes’, ‘Muscle Imbalance’, and ‘Kinetic Control’ books, in which diagnostic theories using movement or musculoskeletal systems were written in relation to musculoskeletal disorders, were selected.^25–27^ Finally, a book called “Therapeutic Exercise,” which describes exercise therapy, one of the options for treating musculoskeletal disorders, was selected.^28^ Document text was extracted using the RecursiveCharacterTextSplitter, and large chapters of the document were divided into small units to increase processing efficiency. The extracted text was embedded by HuggingFaceInstructorEmbedding. The embedded document was then converted into a database format using Chroma to enable saving and retrieval.

**Figure 1.**
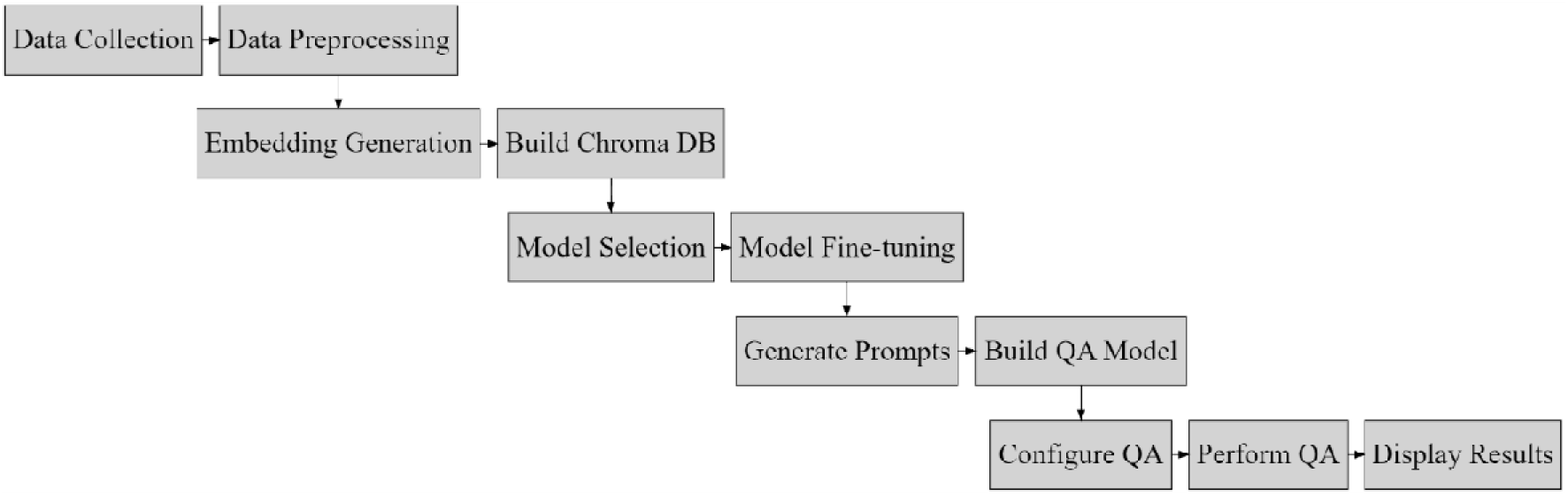
Flowchart of this study.

### Model selection and Fine-tuning

The Llama-2-13B Chat model was chosen to provide advanced features for natural language processing and question-answering tasks. This model is specifically designed for interactive AI and question-answering scenarios. The Lama-2-13B model was loaded using the AutoGPTQForCausalLM library, and the settings required for model fine-tuning were performed. TextStreamer, a component that processes input text, was used to process text data effectively. This step was important in preparing and formatting the input text before feeding it to the model. The process of creating a response using the Lama-2-13B model by constructing the HuggingFacePipeline has been simplified. This pipeline has been applied to generate consistent and contextually relevant responses by managing various aspects such as tokenization and decoding during the creation process. Finally, the RetrivingQA framework was used to develop a QA model with added search functionality. Through this process, a Fine-tuned QA model was established, which is expected to provide more accurate and contextual responses to various musculoskeletal disease diagnosis and treatment-related questions.

### Input Questions and Base Comparison of Models

Questions expected to be addressed primarily in each book were generated, and input data were prepared to communicate those questions to the model. The questions created were entered into the base and fine-tuned models, respectively. The questions created and entered according to the selected books were as shown in table 1.

**Table 1.** Questions related diagnosis and treatment of musculoskeletal system.

| List of Questions |
| --- |
| 1) Tell me about the action of serratus anterior muscle |
| 2) Tell me about the scapulo-humeral rhythm |
| 3) Tell me about the Scapular Downward Rotation movement impairment syndrome |
| 4) Tell me about the upper crossed syndrome |
| 5) Tell me about the kinetic medial rotation test |
| 6) Tell me management for glenohumeral joint after surgery |

## RESULTS

The answers generated for each question of the base and the fine-tuned models are presented in Table 2. The base model often failed to understand or provide relevant information because the questions were unclear or key terms were not recognized. In contrast, the fine-tuned model consistently produced answers specific to the musculoskeletal context, demonstrating improved understanding of specialized topics. In particular, the fine-tuned model did not show any lack of information for any question and showed improved performance in processing musculoskeletal inquiries based on fine-tuned text content. Regarding the serratus anterior muscle, the base model provided a general overview of its role in shoulder movement and stabilization. In contrast, the fine-tuned model provided a more detailed description, emphasizing specific movements during arm lifting and outcomes related to muscle paralysis. For the scapulo-humeral rhythm, the base model was insufficient to provide information due to lack of context, but the fine-tuned model provided a comprehensive definition, including details on the “set-up phase” and the constant rate of humerus and scapula movement during shoulder flexion. Regarding scapular downward rotation syndrome, the base model was unfamiliar and additional information was requested. In contrast, the fine-tuned model not only defined the major movement disorders but also outlined the associated muscle dysfunction, symptoms, and available treatment options. In the case of upper crossed syndrome, the base model acknowledged ignorance of the term and suggested that more context was needed. Conversely, the fine-tuned model identified specific muscle tension and weakness patterns associated with the syndrome, providing a detailed description. Regarding the kinetic medial rotation test, the base model lacked information about the test and provided speculative guesses based on the phrases provided. In contrast, the fine-tuned model describes the components of the test in more detail and explains how the results contribute to assessing rotator cuff muscles. Regarding postoperative management of the glenohumeral joint, the base model provides general postoperative management principles, emphasizing the need for individualized advice. On the other hand, fine-tuned models represent a more nuanced understanding, emphasizing the importance of early mobilization, exercise, and potentially surgical stabilization for specific cases.

**Table 2.**
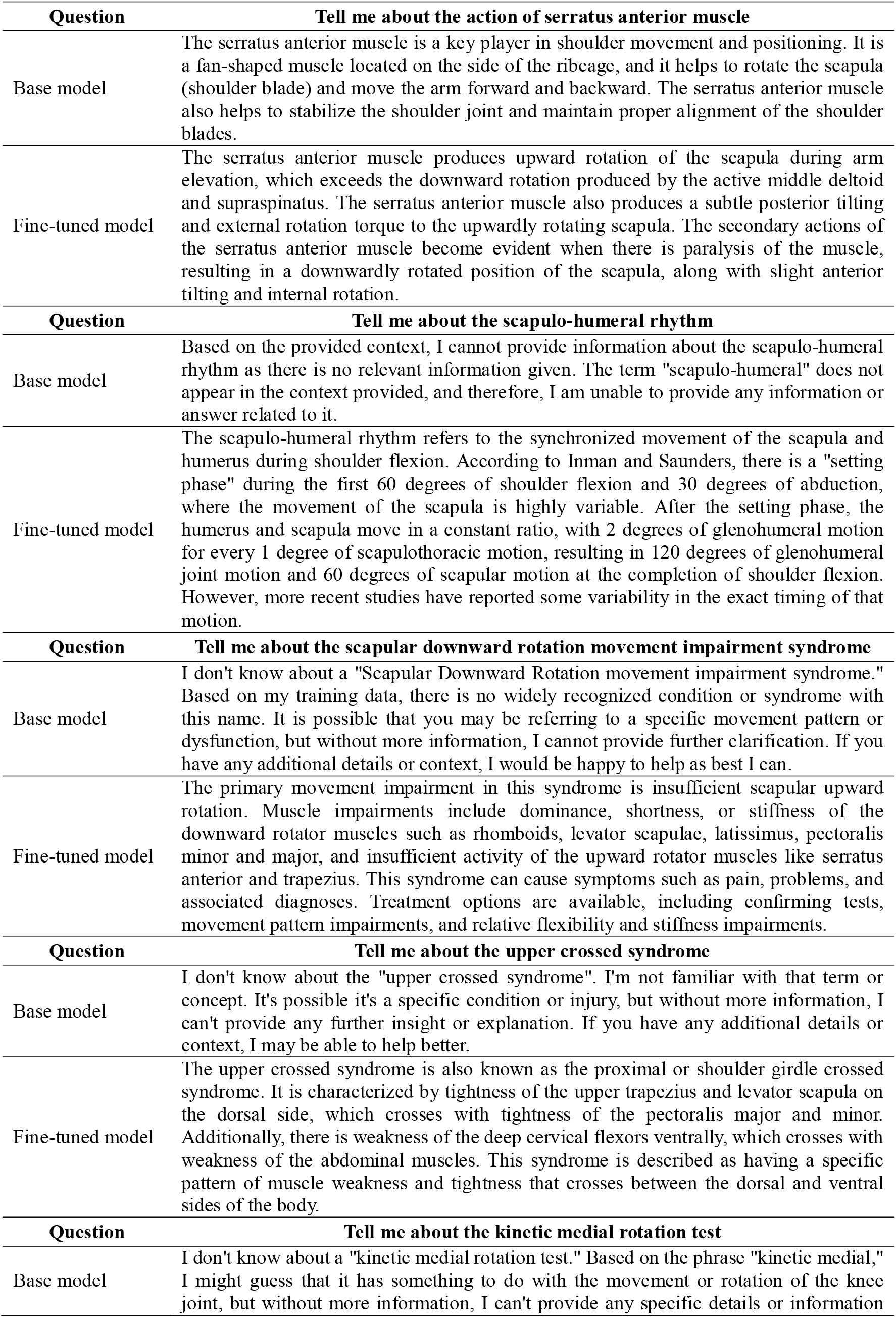
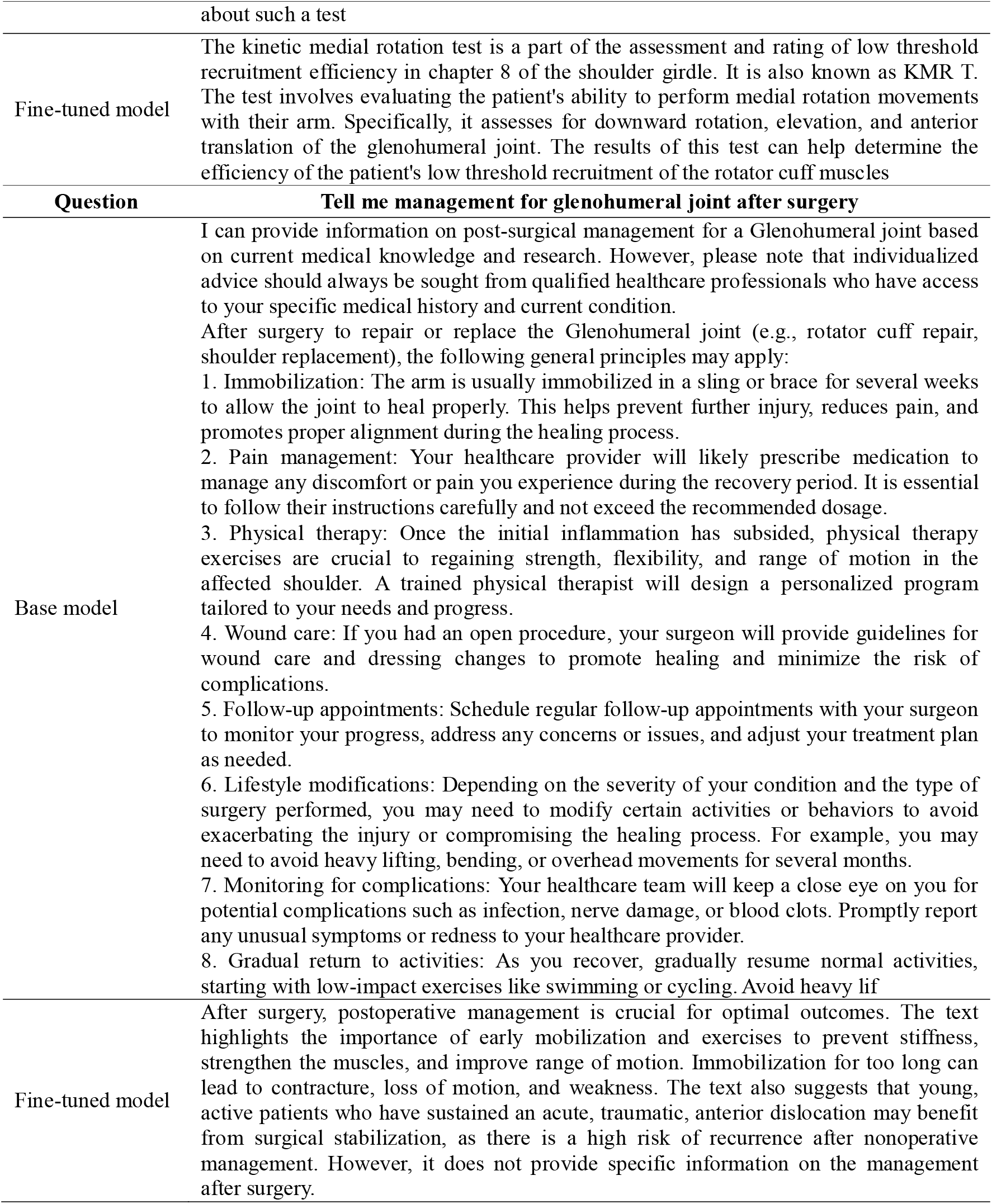
Questions and answers from models before and after fine-tuning.

| Question | Tell me about the action of serratus anterior muscle |
| --- | --- |
| Base model | The serratus anterior muscle is a key player in shoulder movement and positioning. It is a fan-shaped muscle located on the side of the ribcage, and it helps to rotate the scapula (shoulder blade) and move the arm forward and backward. The serratus anterior muscle also helps to stabilize the shoulder joint and maintain proper alignment of the shoulder blades. |
| Fine-tuned model | The serratus anterior muscle produces upward rotation of the scapula during arm elevation, which exceeds the downward rotation produced by the active middle deltoid and supraspinatus. The serratus anterior muscle also produces a subtle posterior tilting and external rotation torque to the upwardly rotating scapula. The secondary actions of the serratus anterior muscle become evident when there is paralysis of the muscle, resulting in a downwardly rotated position of the scapula, along with slight anterior tilting and internal rotation. |
| Question | Tell me about the scapulo-humeral rhythm |
| Base model | Based on the provided context, I cannot provide information about the scapulo-humeral rhythm as there is no relevant information given. The term "scapulo-humeral" does not appear in the context provided, and therefore, I am unable to provide any information or answer related to it. |
| Fine-tuned model | The scapulo-humeral rhythm refers to the synchronized movement of the scapula and humerus during shoulder flexion. According to Inman and Saunders, there is a "setting phase" during the first 60 degrees of shoulder flexion and 30 degrees of abduction, where the movement of the scapula is highly variable. After the setting phase, the humerus and scapula move in a constant ratio, with 2 degrees of glenohumeral motion for every 1 degree of scapulothoracic motion, resulting in 120 degrees of glenohumeral joint motion and 60 degrees of scapular motion at the completion of shoulder flexion. However, more recent studies have reported some variability in the exact timing of that motion. |
| Question | Tell me about the scapular downward rotation movement impairment syndrome |
| Base model | I don't know about a "Scapular Downward Rotation movement impairment syndrome." Based on my training data, there is no widely recognized condition or syndrome with this name. It is possible that you may be referring to a specific movement pattern or dysfunction, but without more information, I cannot provide further clarification. If you have any additional details or context, I would be happy to help as best I can. |
| Fine-tuned model | The primary movement impairment in this syndrome is insufficient scapular upward rotation. Muscle impairments include dominance, shortness, or stiffness of the downward rotator muscles such as rhomboids, levator scapulae, latissimus, pectoralis minor and major, and insufficient activity of the upward rotator muscles like serratus anterior and trapezius. This syndrome can cause symptoms such as pain, problems, and associated diagnoses. Treatment options are available, including confirming tests, movement pattern impairments, and relative flexibility and stiffness impairments. |
| Question | Tell me about the upper crossed syndrome |
| Base model | I don't know about the "upper crossed syndrome". I'm not familiar with that term or concept. It's possible it's a specific condition or injury, but without more information, I can't provide any further insight or explanation. If you have any additional details or context, I may be able to help better. |
| Fine-tuned model | The upper crossed syndrome is also known as the proximal or shoulder girdle crossed syndrome. It is characterized by tightness of the upper trapezius and levator scapula on the dorsal side, which crosses with tightness of the pectoralis major and minor. Additionally, there is weakness of the deep cervical flexors ventrally, which crosses with weakness of the abdominal muscles. This syndrome is described as having a specific pattern of muscle weakness and tightness that crosses between the dorsal and ventral sides of the body. |
| Question | Tell me about the kinetic medial rotation test |
| Base model | I don't know about a "kinetic medial rotation test." Based on the phrase "kinetic medial," I might guess that it has something to do with the movement or rotation of the knee joint, but without more information, I can't provide any specific details or information |
|  | about such a test |
| Fine-tuned model | The kinetic medial rotation test is a part of the assessment and rating of low threshold recruitment efficiency in chapter 8 of the shoulder girdle. It is also known as KMR T. The test involves evaluating the patient's ability to perform medial rotation movements with their arm. Specifically, it assesses for downward rotation, elevation, and anterior translation of the glenohumeral joint. The results of this test can help determine the efficiency of the patient's low threshold recruitment of the rotator cuff muscles |
| **Question** | **Tell me management for glenohumeral joint after surgery** |
| Base model | I can provide information on post-surgical management for a Glenohumeral joint based on current medical knowledge and research. However, please note that individualized advice should always be sought from qualified healthcare professionals who have access to your specific medical history and current condition.<br>After surgery to repair or replace the Glenohumeral joint (e.g., rotator cuff repair, shoulder replacement), the following general principles may apply:<br>1. Immobilization: The arm is usually immobilized in a sling or brace for several weeks to allow the joint to heal properly. This helps prevent further injury, reduces pain, and promotes proper alignment during the healing process.<br>2. Pain management: Your healthcare provider will likely prescribe medication to manage any discomfort or pain you experience during the recovery period. It is essential to follow their instructions carefully and not exceed the recommended dosage.<br>3. Physical therapy: Once the initial inflammation has subsided, physical therapy exercises are crucial to regaining strength, flexibility, and range of motion in the affected shoulder. A trained physical therapist will design a personalized program tailored to your needs and progress.<br>4. Wound care: If you had an open procedure, your surgeon will provide guidelines for wound care and dressing changes to promote healing and minimize the risk of complications.<br>5. Follow-up appointments: Schedule regular follow-up appointments with your surgeon to monitor your progress, address any concerns or issues, and adjust your treatment plan as needed.<br>6. Lifestyle modifications: Depending on the severity of your condition and the type of surgery performed, you may need to modify certain activities or behaviors to avoid exacerbating the injury or compromising the healing process. For example, you may need to avoid heavy lifting, bending, or overhead movements for several months.<br>7. Monitoring for complications: Your healthcare team will keep a close eye on you for potential complications such as infection, nerve damage, or blood clots. Promptly report any unusual symptoms or redness to your healthcare provider.<br>8. Gradual return to activities: As you recover, gradually resume normal activities, starting with low-impact exercises like swimming or cycling. Avoid heavy lif |
| Fine-tuned model | After surgery, postoperative management is crucial for optimal outcomes. The text highlights the importance of early mobilization and exercises to prevent stiffness, strengthen the muscles, and improve range of motion. Immobilization for too long can lead to contracture, loss of motion, and weakness. The text also suggests that young, active patients who have sustained an acute, traumatic, anterior dislocation may benefit from surgical stabilization, as there is a high risk of recurrence after nonoperative management. However, it does not provide specific information on the management after surgery. |

## DISCUSSION

The base model generally provided accurate information, but it lacked detailed information about certain terms or concepts. However, general information was provided on postoperative management of the glenohumeral joint. The fine-tuned model provided more detailed and specialized information on specific terms and concepts. It may provide more contextual information, but there may be differences in the quality of the response. It was suggested that fine-tuned models tend to provide more detailed and context-specific answers, especially in areas related to specific muscle activities, syndromes, and tests. For serratus anterior, scapula-humeral rhythm, scapular inferior rotation syndrome, superior crossed syndrome, and kinetic medial rotation test, fine-tuned model provided in-depth insights, whereas base model often provided uninformed or speculative guesses. Overall, the fine-tuned model demonstrated superior performance, demonstrating its ability to provide more detailed and contextual information across a variety of musculoskeletal topics compared to the base model, which provides general information.

These results were consistent with various studies showing that model tuning enables effective and expert responses to questions by equipping models with specific knowledge of specialized domains.^6,11,14^ Several studies have already attempted to fine-tune large-scale language models or build medical language models in the medical field.^19,29,30^ Yang et al (2019) introduced a large-scale clinical language model called GatorTron, developed with more than 9 billion words of text, and the generative model using this language model led to the improvement of medical question answers.^29^ Additionally, Lu et al. (2022) introduced the ClinicalT5 model and said that this model can be applied to various tasks in the medical field because it can understand and process medical terminology, context, and special language structures.^30^ Nova (2023) suggested that using a GLM learned from text data such as medical records or from medical personnel’s voice data could simplify and provide medical information that was difficult for patients to understand.^19^

Development of a language model in a specific field such as the above or improvement of the model through fine-tuning is also used for education in that field.^8,11,21^ Karabacak et al. (2023) suggested that AI and GLMs will provide significant opportunities to improve medical education through realistic simulations, digital patients, personalized feedback, assessment methods, and removal of language barriers.^21^ Unlike previous studies that used medical records, this study used text data from books containing knowledge about the musculoskeletal system in the field of physical therapy to fine-tune the model, but it is expected to be helpful in education as well. This is an important advancement that could provide a new dimension in expanding medical education and expertise. This approach is expected to contain specific knowledge about the field and provide more detailed information, allowing medical professionals and educators to provide learners more effectively with an in-depth understanding of specific topics. The expertise gained through fine-tuning the model not only increases its applicability in clinical situations, but also provides learners with rich experience of real-world situations in the field. This realistic training environment can improve the practical skills of healthcare professionals and increase the effectiveness of research-based training. Additionally, the utilization of GLM can overcome language barriers and improve the quality of global medical education. Able to understand and handle multiple languages and cultures, GLM can foster international educational collaboration and improve information exchange between healthcare professionals.

However, several problems are being raised in content creation using AI and GLM. Among them are content quality, bias, and ethical and legal issues.^21^ In addition, it is required to develop guidelines and policies in evaluating the accuracy of AI-generated content.^20,21^ To this end, it is said that reliability and reliability can be increased by sharing information on data and evaluation methods used in education.^21^ Continuous research and academic cooperation are needed to realize the maximum potential of AI and GLM in medical education and to respond to potential risks and challenges at the same time.^21^ It is said that through the cooperation of medical experts, these technologies will be integrated in an effective and responsible way, leading to improved medical care between learning experiences and patients.^20,21^

There were some limitations to this study. First, it consists of books that focus on specific topics such as musculoskeletal diagnosis and treatment, which may prevent the model from being exposed to information about various patient cases or medical conditions and thus may not provide general information. In addition, certain medical books may be biased in their diagnosis and treatment depending on the specific medical approach in their diagnosis and treatment methods. Finally, if the model is updated only with information from medical books, it may not always reflect empirical knowledge or up-to-date medical information about diagnosis and treatment.

## CONCLUSION

The model fine-tuned with books on musculoskeletal diagnosis and treatment showed superior performance in providing various information related to the topic compared to the basic model. This demonstrates the effectiveness of model tuning for a specific knowledge domain and suggests that model enhancement using textual data on the musculoskeletal system may help in medical education and expertise acquisition. However, there will be a need for guidelines and policies to evaluate reliable AI-generated content, and there will be a need to increase reliability by sharing information on data and evaluation methods used in education.

## Data Availability

All data produced in the present study are available upon reasonable request to the authors

